# Liver Function in Novel Coronavirus Disease (COVID-19): A Systematic Review and Meta-Analysis

**DOI:** 10.1101/2020.05.20.20108357

**Authors:** Mohammad Zahedi, Mohammad Yousefi, Mahdi Abounoori, Mohammad Malekan, Fatemeh Tajik, Keyvan Heydari, Parham Mortazavi, Monireh Ghazaeian, Fateme Sheydaee, Amirreza Nasirzadeh, Reza Alizadeh-Navaei

## Abstract

**Introduction:** The outbreak of new coronavirus has become a global public health challenge. Given a consequential liver function, and the high risk of death coming from liver disorders, the assessment of Novel Coronavirus Disease on liver function is importance. Hence, we carried out this meta-analysis to heightening insight into the occult features of COVID 19, which is likely to affect liver function.

**Method:** This study was performed using databases of Web of Science, Scopus, and PubMed. We considered English cross-sectional and case-series papers, which reported available findings on the association between liver injury and COVID-19 infection. We used the STATA v.11 and random effect model for data analysis.

**Result:** In this present meta-analysis, 52 papers, including 8,463 COVID-19 patients, were studied. The prevalence of increased liver enzymes among the patients, including Alanine aminotransferase, Aspartate aminotransferase, were 30% and 21% in non-severe patients, respectively, which were 38% and 48% in severe patients. The prevalence of increasing C-reactive protein, Lactate dehydrogenase, D-dimer, and Bilirubin were 55%, 39%, 28%, and 10% in non-severe patients respectively, which were 78%, 75%, 79% and 17% in sever patients.The prevalence of liver toxicity as a complication of COVID-19 was 20%.Also patients who have severe condition are 5.54, 4.22, 4.96, 4.13 and 4.34 times more likely to have elevated CRP, ALT, AST, LDH, D-dimer enzymes retrospectively.

**Conclusion:** Elevation of some liver markers were higher in patients with severe COVID-19 infection. All to gather, we assumed that abnormal liver markers could act as a prognostic factor for a better survey of COVID-19.

## Introduction

The outbreak of a new virus, called the 2019 novel coronavirus (2019 n-COV), has been led to many worries amid all strata of populations, especially the vast majority of health care workers (1). By 11 May 2020, a considerable proportion of the population, approximately 4006257, was detected as Coronavirus cases, and the rate of death Toll, relatively 278892, reported. The rate of mortality and new cases highly progress especially, among the vast spectrum of the population comprised of older people (2). 2019 n-COV, in the family coronaviridae, are enveloped, single-stranded positive RNA viruses with different diameters from 60 to 140 nm (3) entered into target cells by binding spike (S) protein to angiotensin-converting enzyme 2 (ACE2) receptor (4). Transmission of this virus is human to human through close contacts or droplets and also feces (5, 6). Health care workers fundamentally deal with the assortment of clinical characteristics that appeared among different patients. Many people are either asymptomatic or mildly symptomatic. The most common clinical features of symptomatic patients consisting of cough, fever, expectoration, myalgia or fatigue, malaise, sore throat, headache, diarrhea, and rarely hemoptysis and vomiting. And also, in severe cases shows a set of shortness of breath and dyspnea (7-12). Some infected patients would suffer from liver dysfunction. Based on a series of studies working on this issue, the changes in liver enzymes among patients, particularly in severe cases, were revealed (13-17). Given a consequential liver function, and the high risk of death coming from liver disorders, the assessment of Novel Coronavirus Disease (COVID-19) on liver function is of significant importance among health care workers (17-19). Also through a recent study on liver enzyme changes during COVID-19 this fact that higher levels of Aspartate aminotransferase (AST), and direct Bilirubin increases the transfer risk to intensive and critical care units, have been stated. It is reported that the AST level of 30.5 (U/L) had a sensitivity of 71.4% and specificity of 68.5% for critical and intensive care transfer (20). Moreover, In a similar study with us, elevated Serum levels of Aspartate aminotransferase, Alanin aminotransferase (ALT), total Bilirubin, and lower levels of Albumin were observed in severe cases (21).

Recent studies on liver function during COVID-19 were not comprehensive, and liver function was not compared with the severity of the disease. This research has reviewed the epistemological and evidential resources on the correlation between COVID-19 and the function of the liver. Hence, we carried out this systematic review and meta-analysis to heightening insight into the occult features of COVID 19, which is likely to affect liver function.

## Method

### Search strategy

The results and their analysis in this study were reported according to the Preferred Reporting Items for Systematic Reviews and Meta-Analyses (PRISMA). We conducted a systematic review and meta-analysis of cross-sectional and case-series studies. Search engines and databases, including Web of Science, Scopus, and PubMed without any time limitation for publications up to April 19, 2020. As a manual search, the list of imported references, a list of related reviews, and the results of Google Scholar have been investigated. The search process was done by two researchers using the following medical subject headings (22) keywords: “2019 novel coronavirus infection” OR COVID19 OR COVID-19 OR “coronavirus disease 2019” OR “coronavirus disease-19” OR “2019-nCoV disease” OR “2019 novel coronavirus disease” OR “2019-nCoV infection” OR “SARS COV-2” OR SARS-COV-2, in combination with “Liver function” OR AST OR ALT OR “Liver toxicity” OR Bilirubin OR INR OR SGOT OR “Aspartate transaminase” OR “Alanine transaminase” OR SGPT OR ALP OR AFP OR liver OR hepat*, which were also combined with and/or/not to find more articles. As a manual search, the list of imported references, list of related reviews, have been investigated.

### Eligibility Criteria

We considered the following criteria for study selection:

1. The study should have an observatory approach investigated COVID-19 patients;
2. Studies on a particular group of people (e.g., pregnant women) has been excluded;
3. Studies included with assessing the association between serum levels of Alanine aminotransferase (ALT), Aspartate aminotransferase (AST), Albumin, Bilirubin, C-Reactive protein, ESR, D-dimer, LDH and severe outcome from COVID-19 infection as the major outcomes of interest and reported median (IQR) for serum levels of AST, ALT, Albumin, Bilirubin in both severe and non-severe COVID-19 infected patients.
4. Studies should be in English. English abstracts of other language studies were investigated for eligible data.

Studies that had incomplete data for any reason and all the case reports, expert opinion articles, review articles, books, and animal studies were excluded.

### Study Selection

Duplicated papers were deleted using EndNote software (version X8, Thomson Reuters, Philadelphia, USA). Two of the authors reviewed the remaining papers according to the criteria, separately. Titles and abstracts were screened for initial study inclusion, with a full-text review when the abstract was insufficient to determine if the study encountered the inclusion or exclusion criteria. In case of disagreement between the two investigators, a third person was making the final decision.

### Quality assessment

For the quality assessment, two members of our team independently valued the selected articles using the modified version of Newcastle-Ottawa Quality Assessment (NOS) and NIH quality assessment tool for case series. The investigated papers categorized into three categories. The studies with a score of 1 or 2 as poor quality, 3 to 6 as moderate quality and studies with a score of 7 to 9 as high quality.

### Data Extraction

Data such as author’s information, publication year, study design, sample size, average/median age and gender of patients, serum levels of CRP, LDH, D-dimer, ESR, AST, ALT, Albumin, and Bilirubin have been extracted and recorded.

### Statistical Analysis

Statistical analysis was performed using STATA v.11 software. The heterogeneity of studies has been investigated using the I-square (I2) test. Based on the results, for I2 more than 50%, we used a random-effects model to pool the results. Moreover, to study the heterogeneity of the added study, subgroup analysis in severe and non-severe patients has been done.

## Results

### Study Selection Process

The search in three databases, including Web of Science, Scopus, and PubMed yields 351 results, and for other sources like google, scholar yields 643 results. After excluding 341 duplicated papers, 663 records were admitted to the screening step. Then, 92 papers have been selected for full-text eligibility assessment. Finally, 58 studies were entered into the meta-analysis. Also, ten studies from other sources were included in the meta-analysis. The PRISMA folw diagram for the study selection process presented in Figure 1.

**Figure 1.**
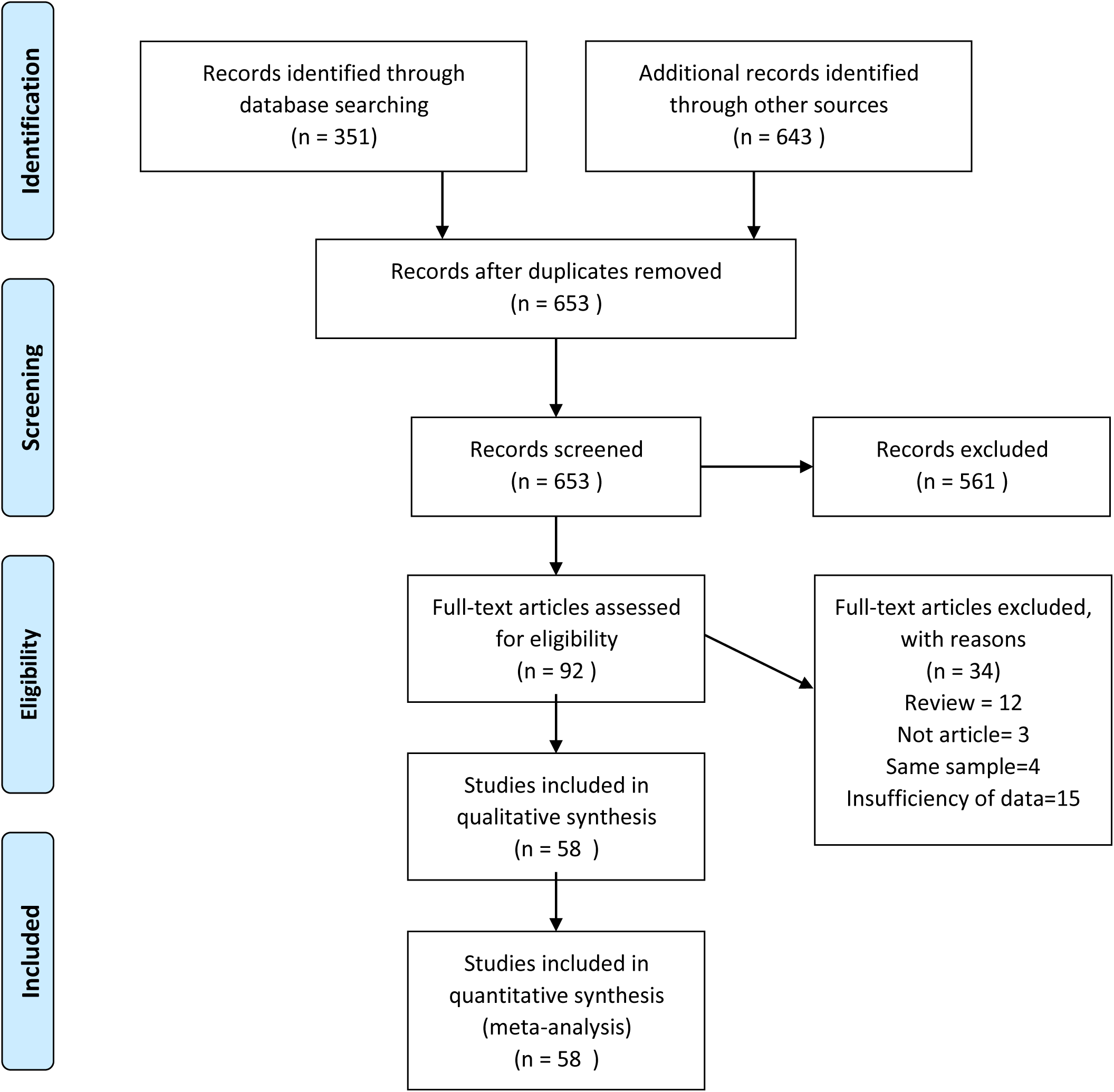
PRISMA flowchart for study selection process

### Study Characteristics

Among selected papers, a total of 8463 patients infected with CoVID-19 with age ranged between1.5 to 90 years o--ld were included in our investigation, and all of the studies were conducted in China. Characteristics of studies entered into meta-analysis are presented in Table 1.

**Table 1.**
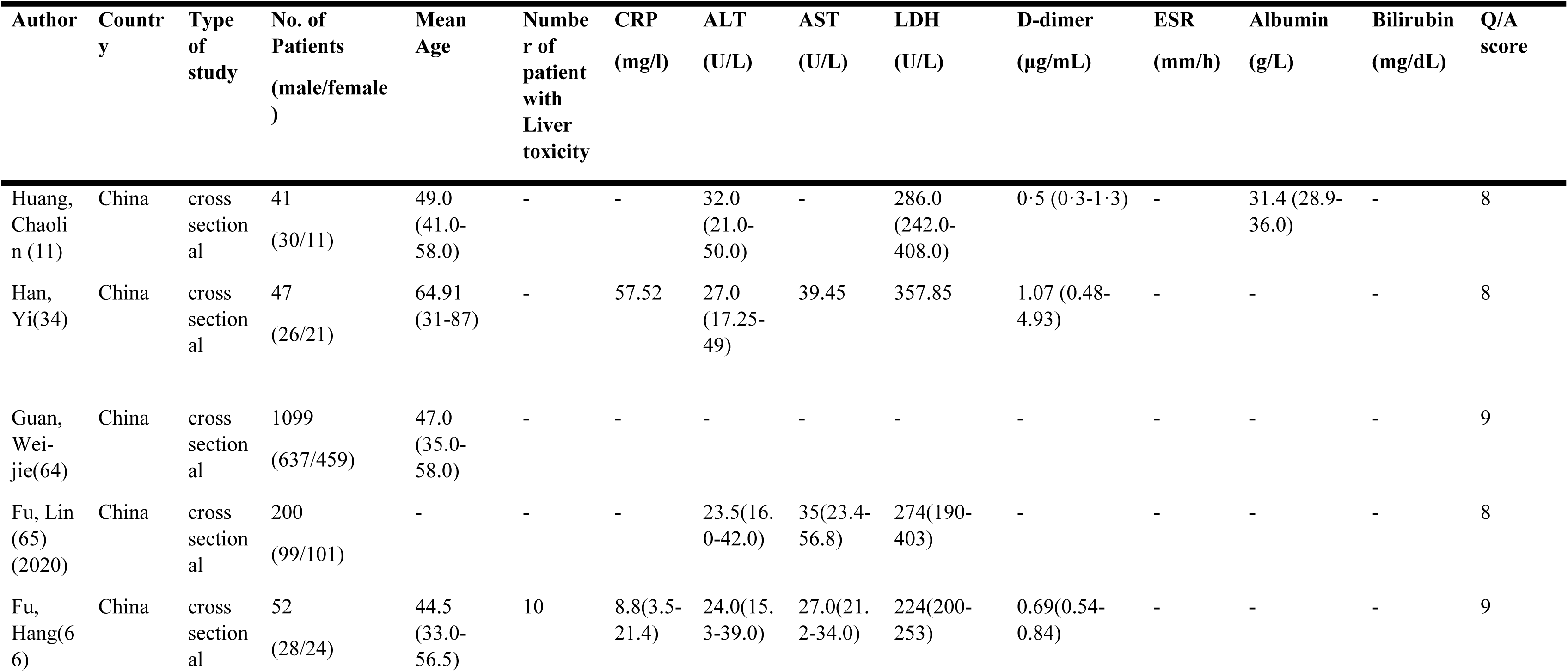
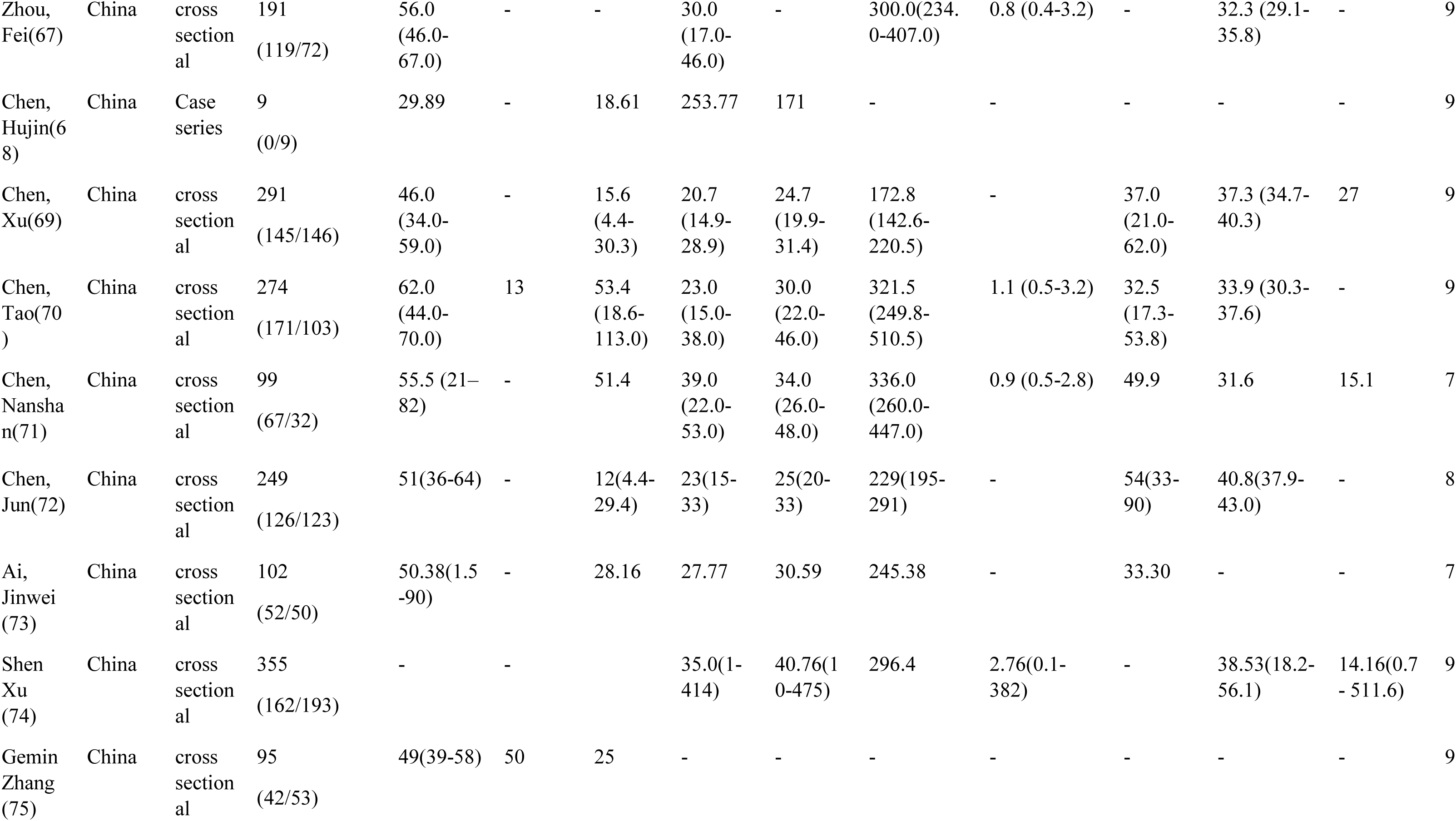
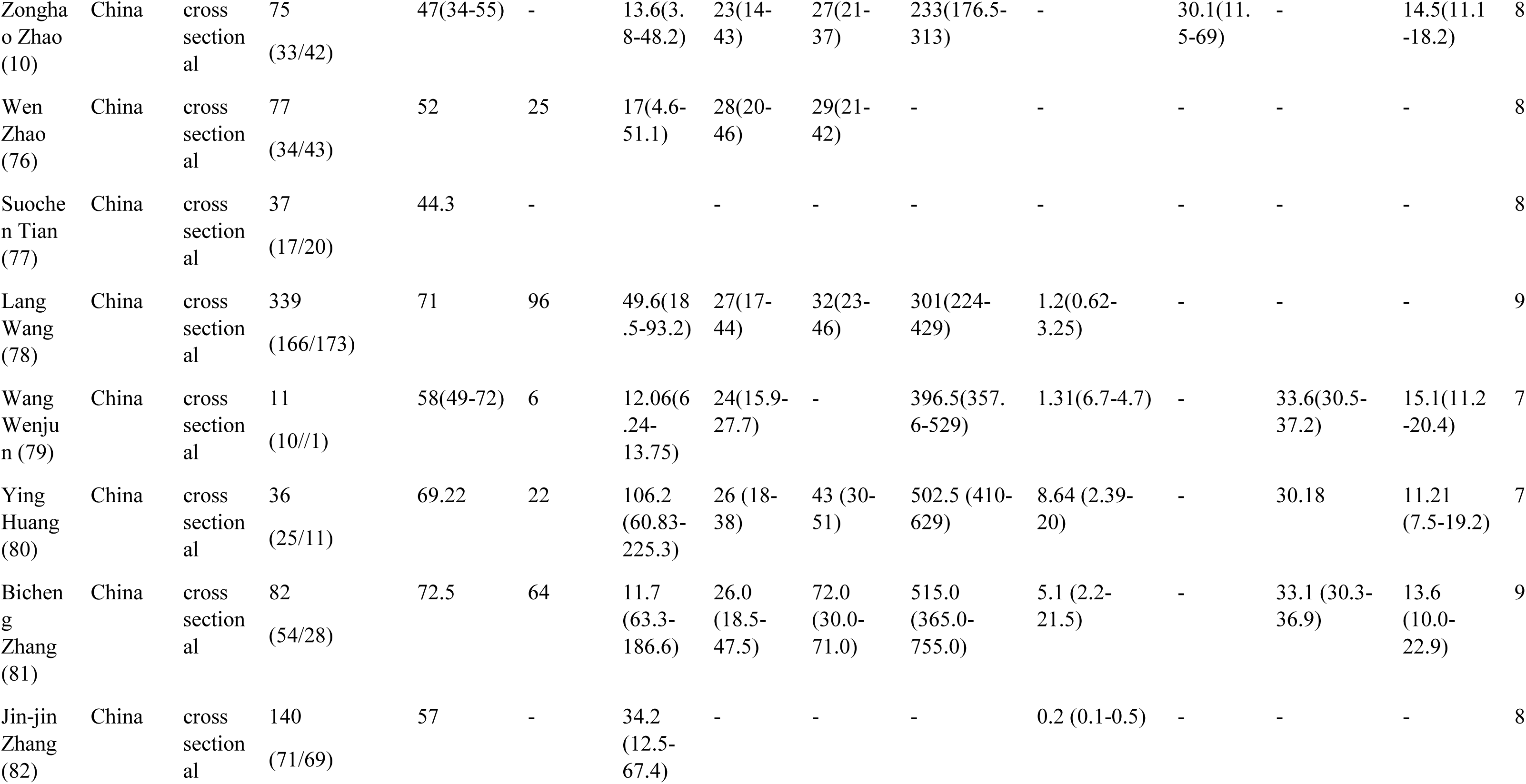
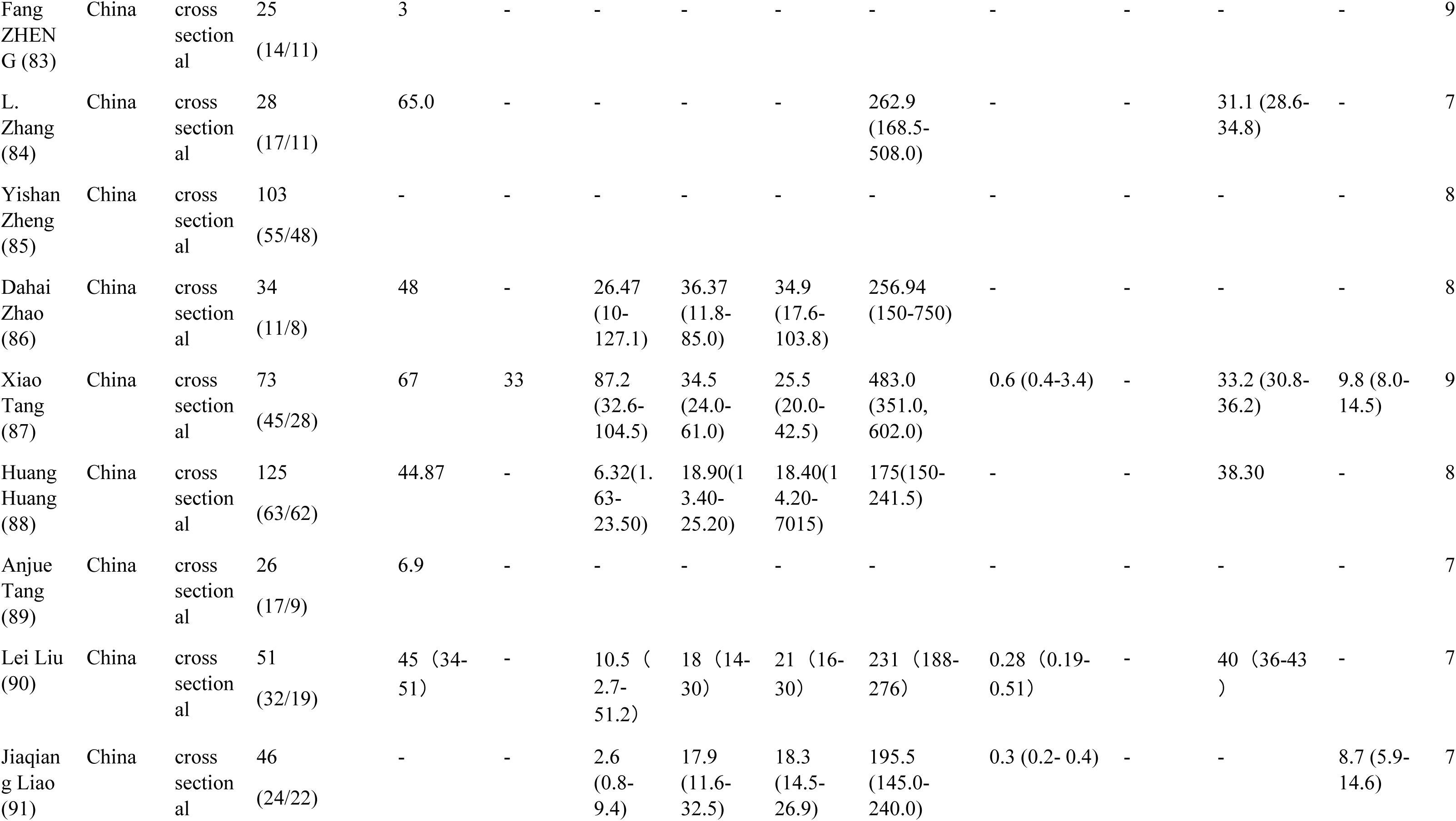
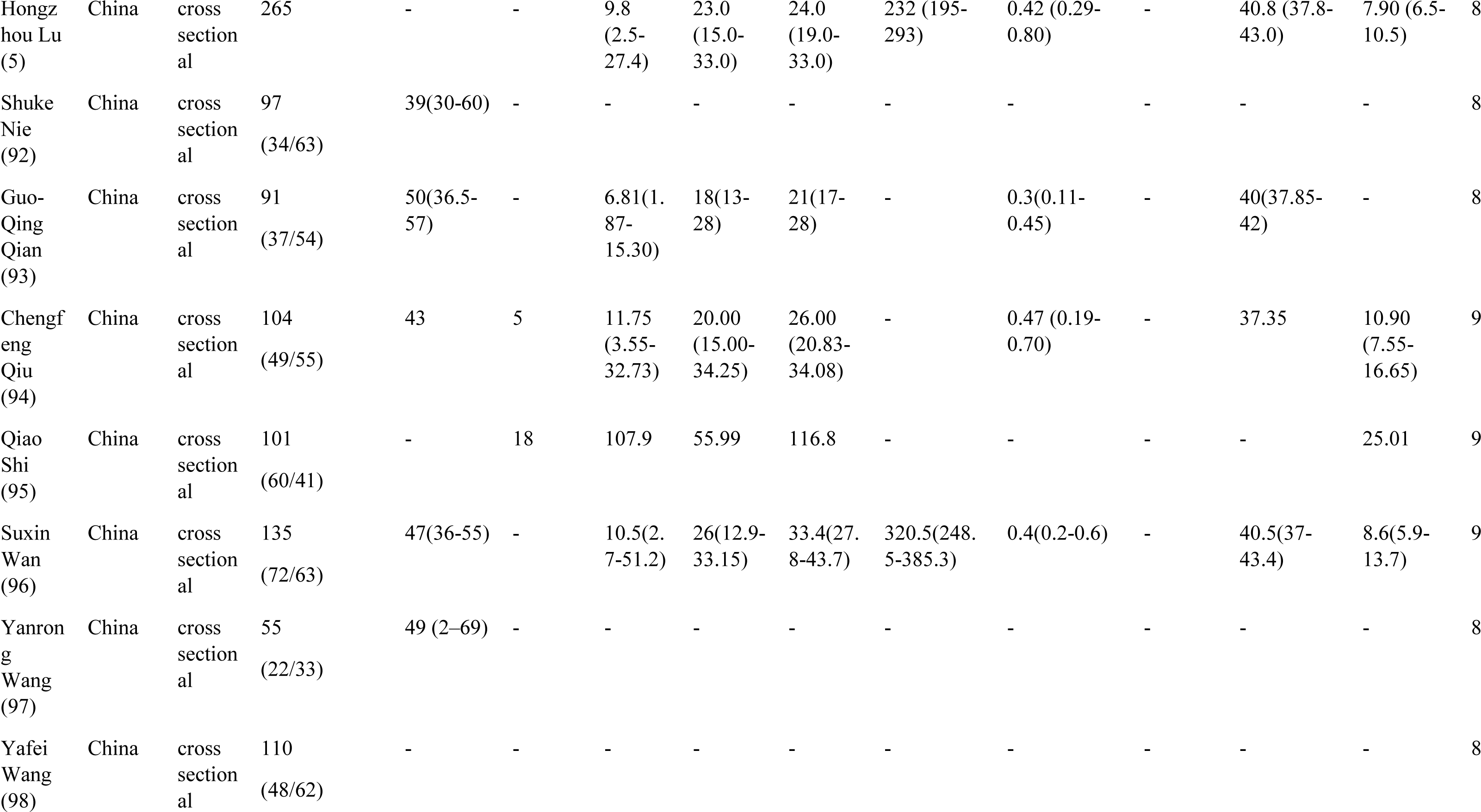
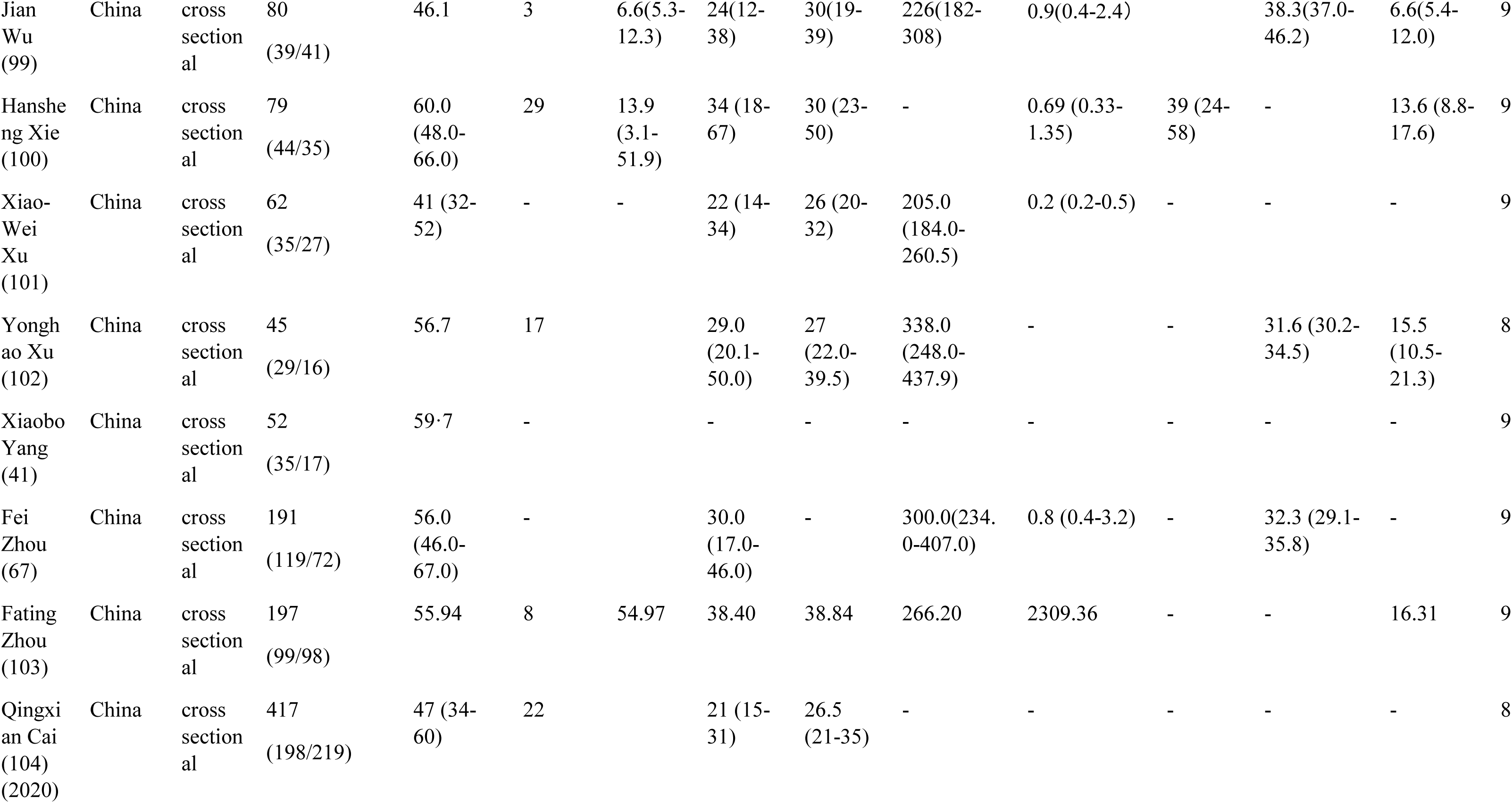
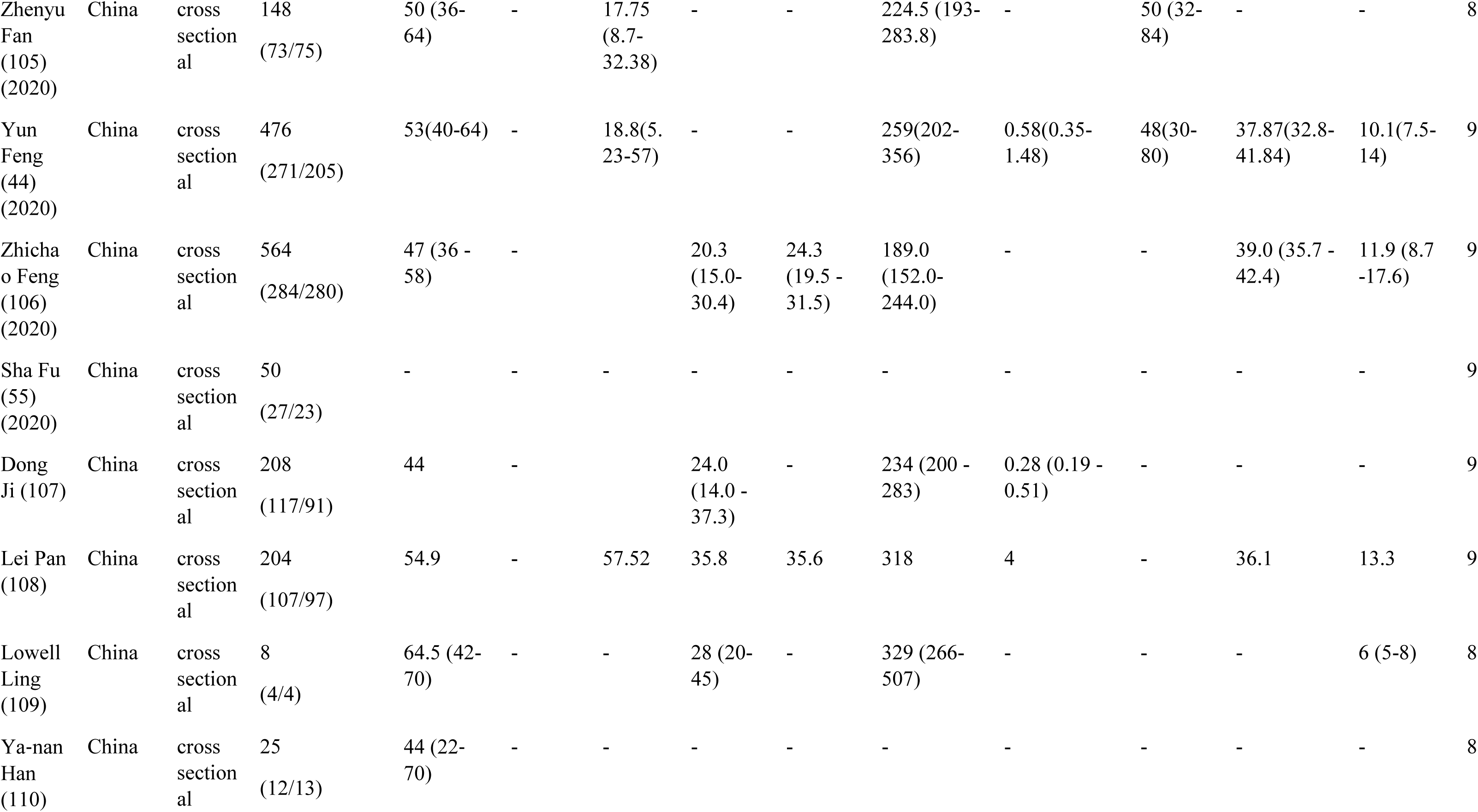
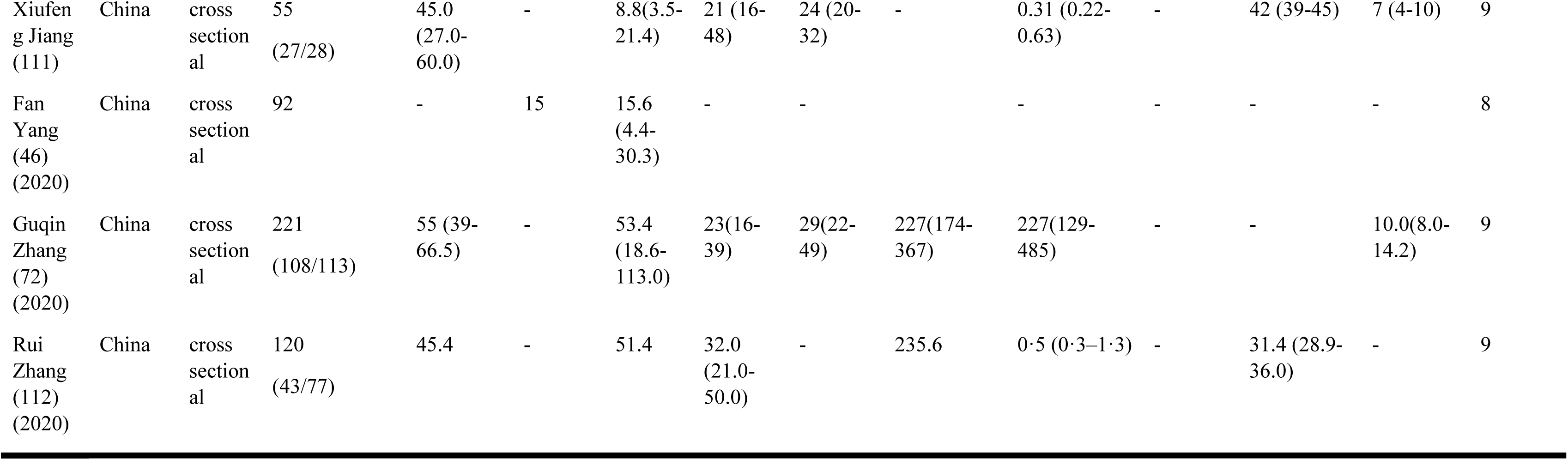
Characteristics of studies entered into the meta-analysis

### Quality Assessment

For quality assessment, the NOS tool for cross-sectional studies and NIH quality assessment tools for case series were conducted.

### Characteristics of Patients

Findings showed that 80% of the patients needed inpatient care services, and 27% were admitted to the Intensive care unit. (Fig. 3) About 50% of patients were discharged; however, 16% of patients were expired (Fig. 4)

**Figure 2.**
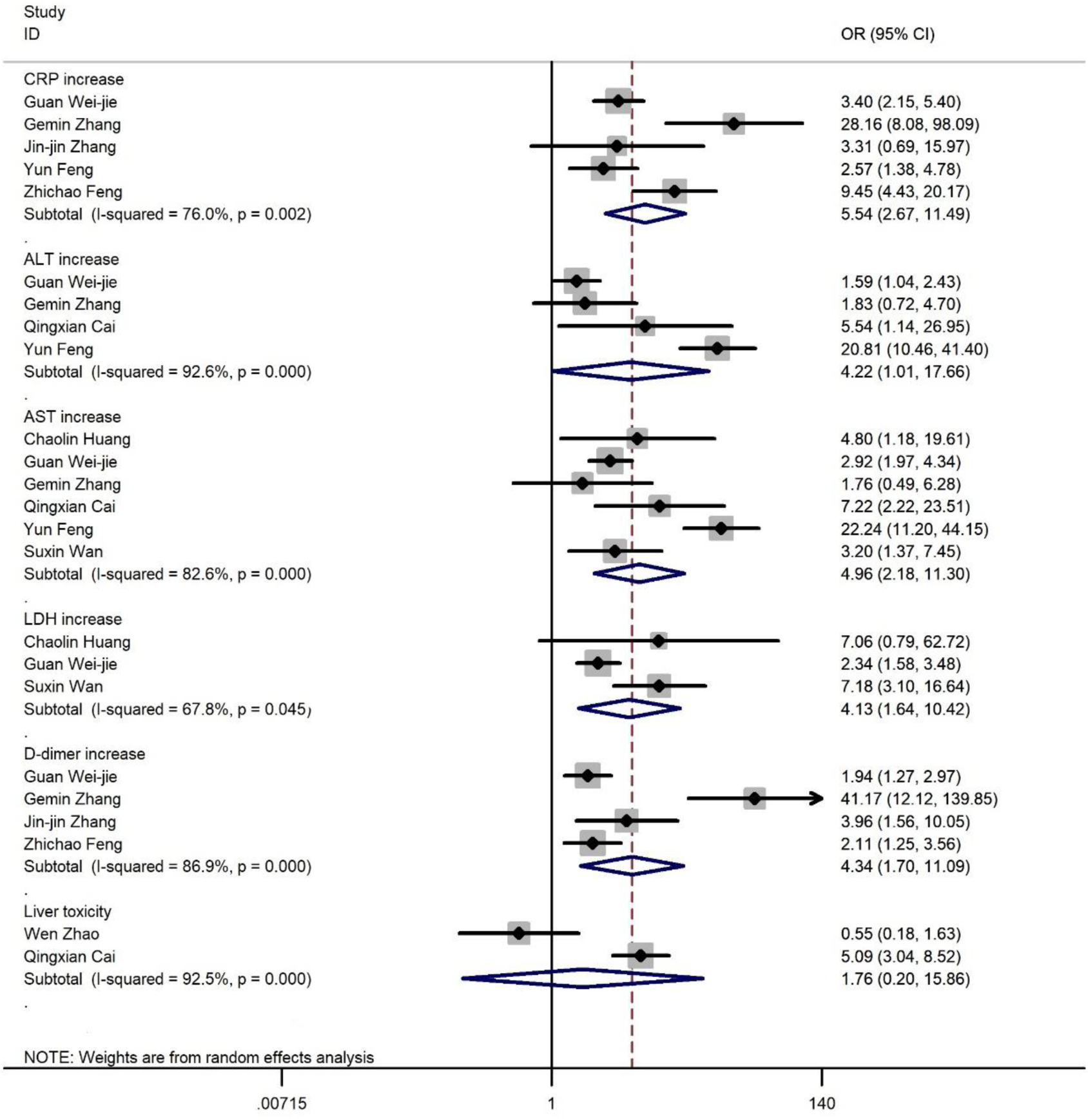
Forest plot for odds ratios of laboratory findings

**Figure 3.**
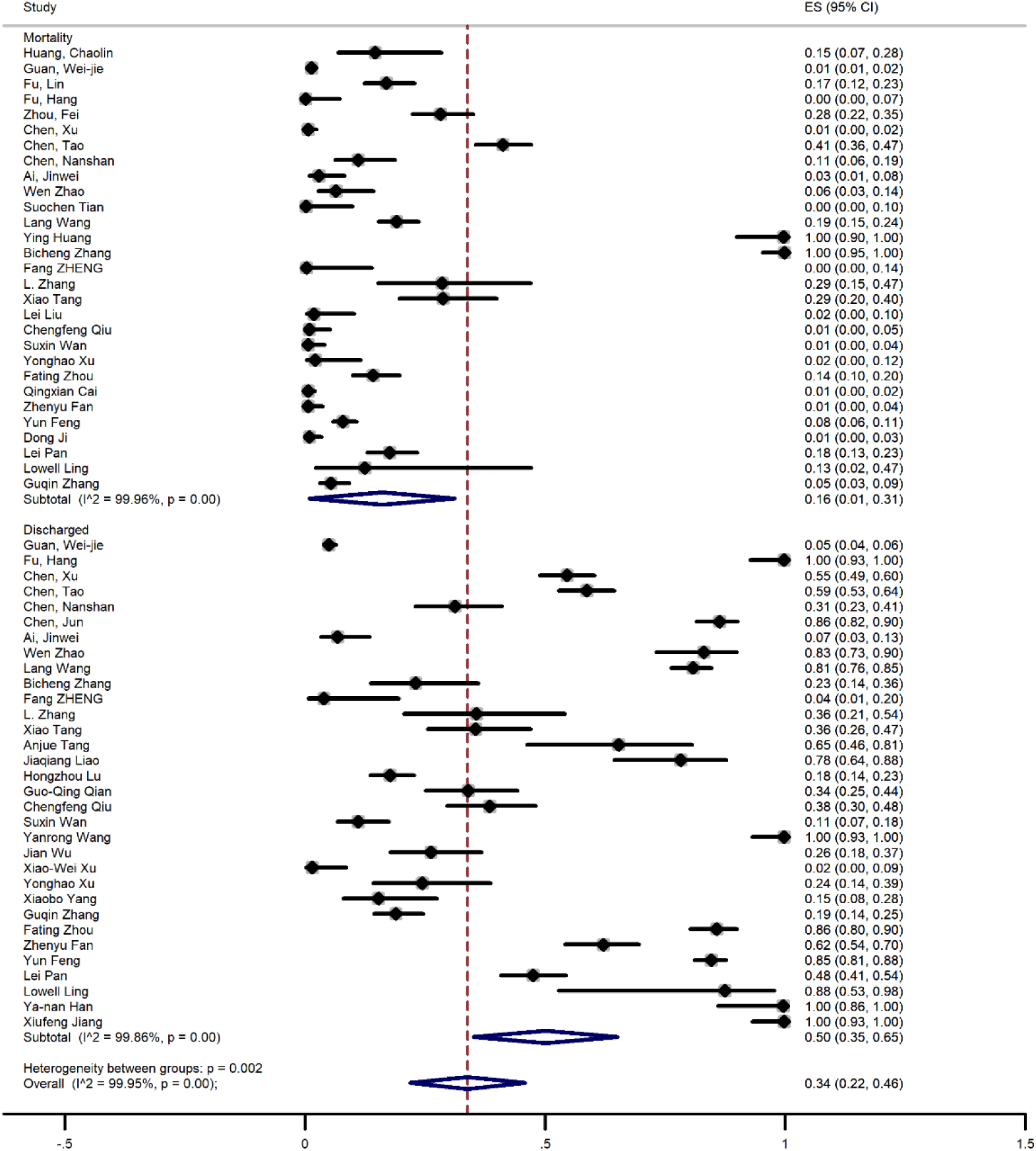
Meta-analysis of prevelance of clinical outcome of patients (A)

**Figure 4.**
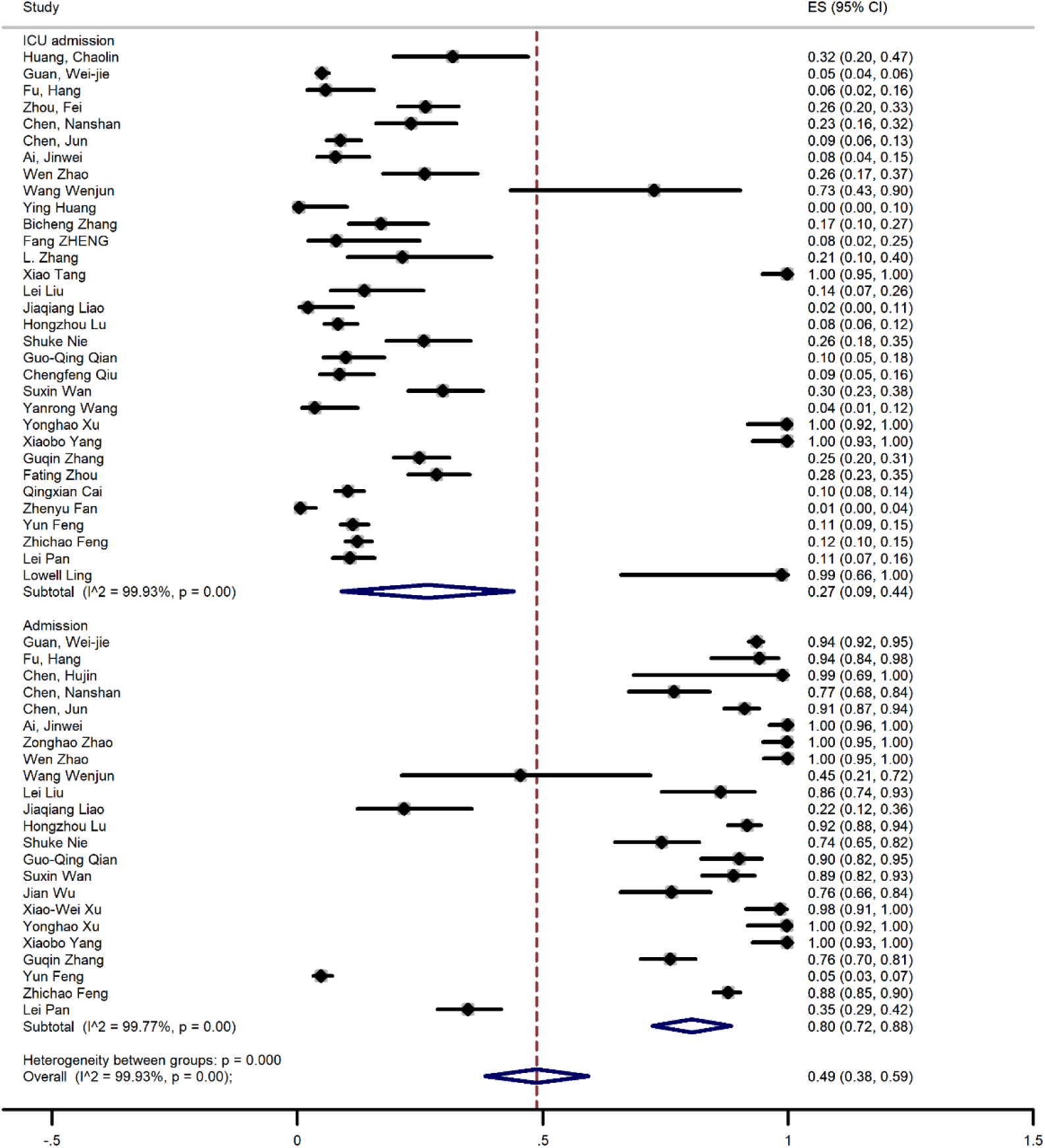
Meta-analysis of prevelance of clinical outcome of patients (B)

### Laboratory Findings

Regarding laboratory findings in patients infected with COVID-19 most common observation was increasing erythrocyte sedimentation rate (ESR), C-reactive protein (CRP), and Lactate dehydrogenase (LDH) which were reported in 9, 18 and 18 studies. Meta-analysis findings are as follows respectively: 57% (95% Cl, 44-70), 53% (95% Cl, 43-62) and 36% (95% Cl, 25-47). (Fig. 5, and Fig. 6).

**Figure 5.**
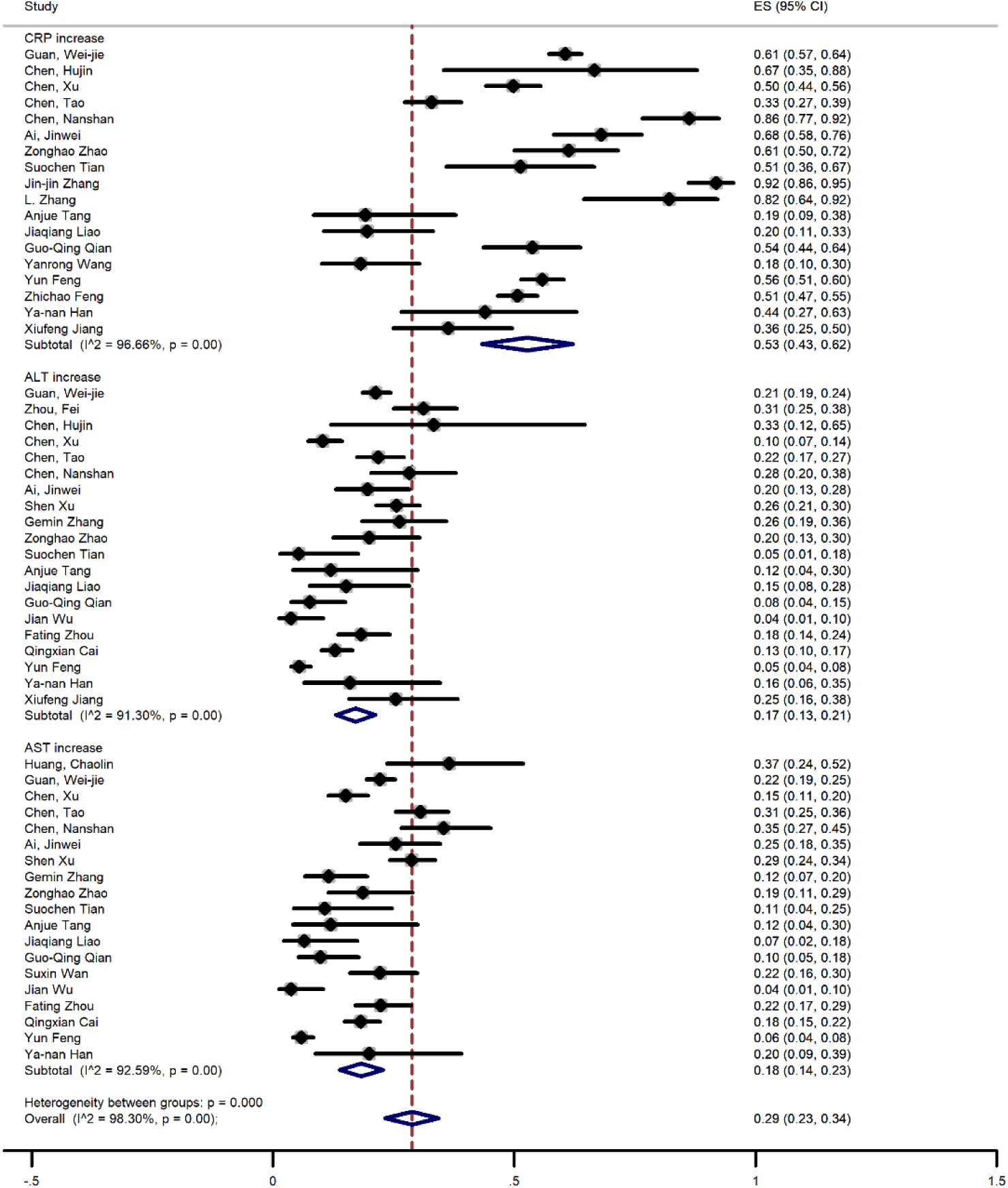
Meta-analysis of prevalence of laboratory findings of all patients (A)

**Figure 6.**
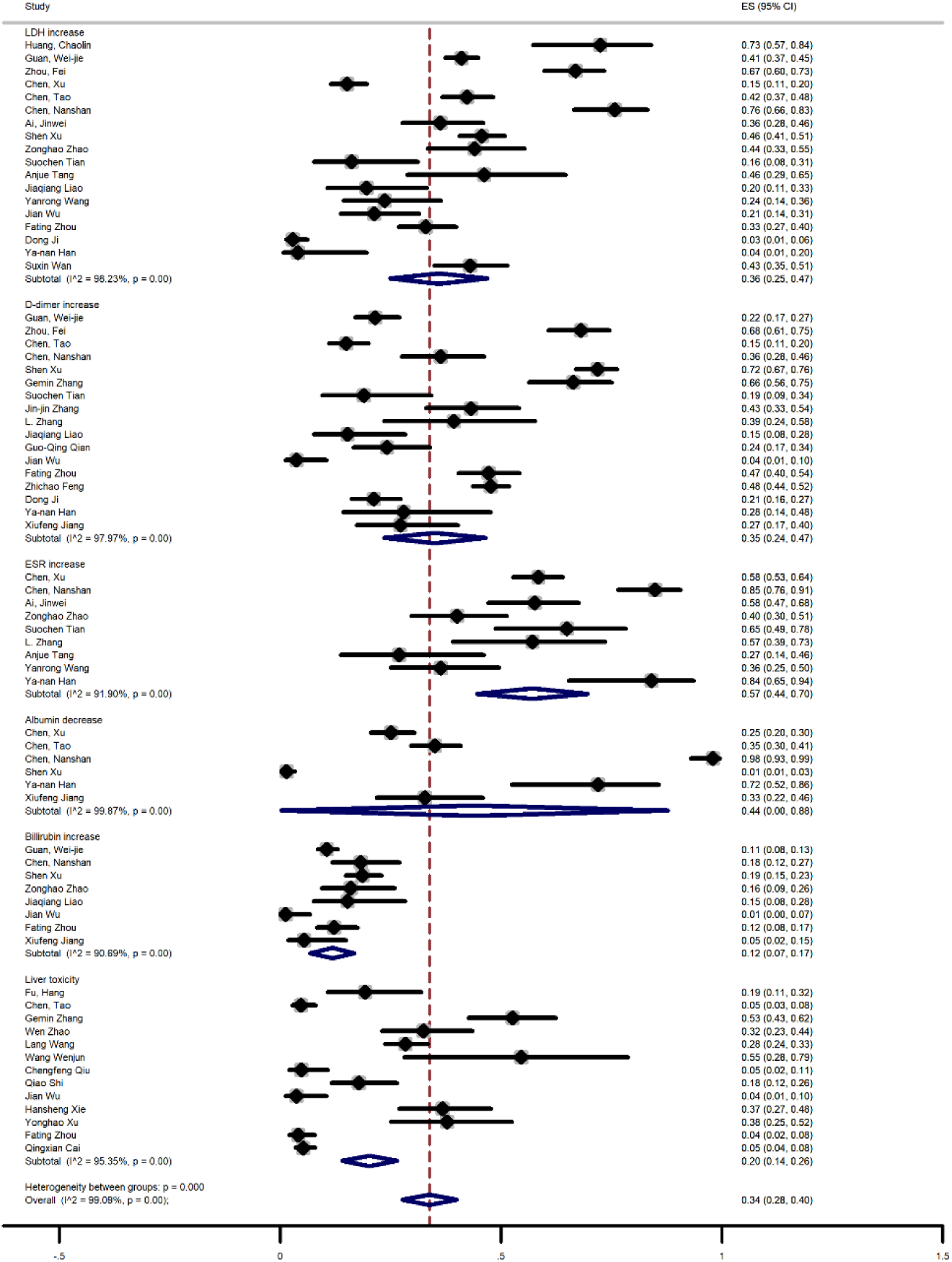
Meta-analysis of prevalence of laboratory findings of all patients (B)

Alanine aminotransferase (ALT), Aspartate aminotransferase (AST), D-dimer, total Bilirubin increasing levels in patients were reported in 20, 19, 17, and 8 studies with the prevalence of 17% (95% Cl, 13-21), 18% (95% Cl, 14-23), 35% (95% Cl, 24-47), and 12% (95% Cl, 7-17) (Fig. 5 and Fig 6). Also decreasing of Albumin level were investigated in 6 studies with prevalence of (44%, 95% Cl, 0-88).

Figure 2 shows the risk estimates results from the random-effect model combining the odds ratios (ORs) for laboratory findings between covid_19 severe and non-severe patients. CRP increase (OR= 5.54, 95% CI= 2.67-11.49, p=0.002), ALT increase (OR= 4.22, 95% CI= 1.01-17.66, p=0.00), AST increase (OR= 4.96, 95% CI= 2.18-11.3, p=0.00), LDH increase (OR= 4.13, 95% CI= 1.64-10.42, p=0.045), D-dimer increase (OR= 4.34, 95% CI= 1.7-11.09, p=0.00), Liver toxicity (OR= 1.67, 95% CI= 0.20-15.86, p=0.00)

### Comorbidities

Different comorbidities were investigated in patients with COVID-19. Hypertension, diabetes, and cardiovascular disease were the most prevalent comorbidities among patients with a prevalence of 24% (95% Cl, 19-29), 15% (95% Cl, 11-18) and 10% (95% Cl, 8-12) that were investigated in 34,35 and 32 studies respectively (Fig.7, Fig.8 and Table 2).

**Figure 7.**
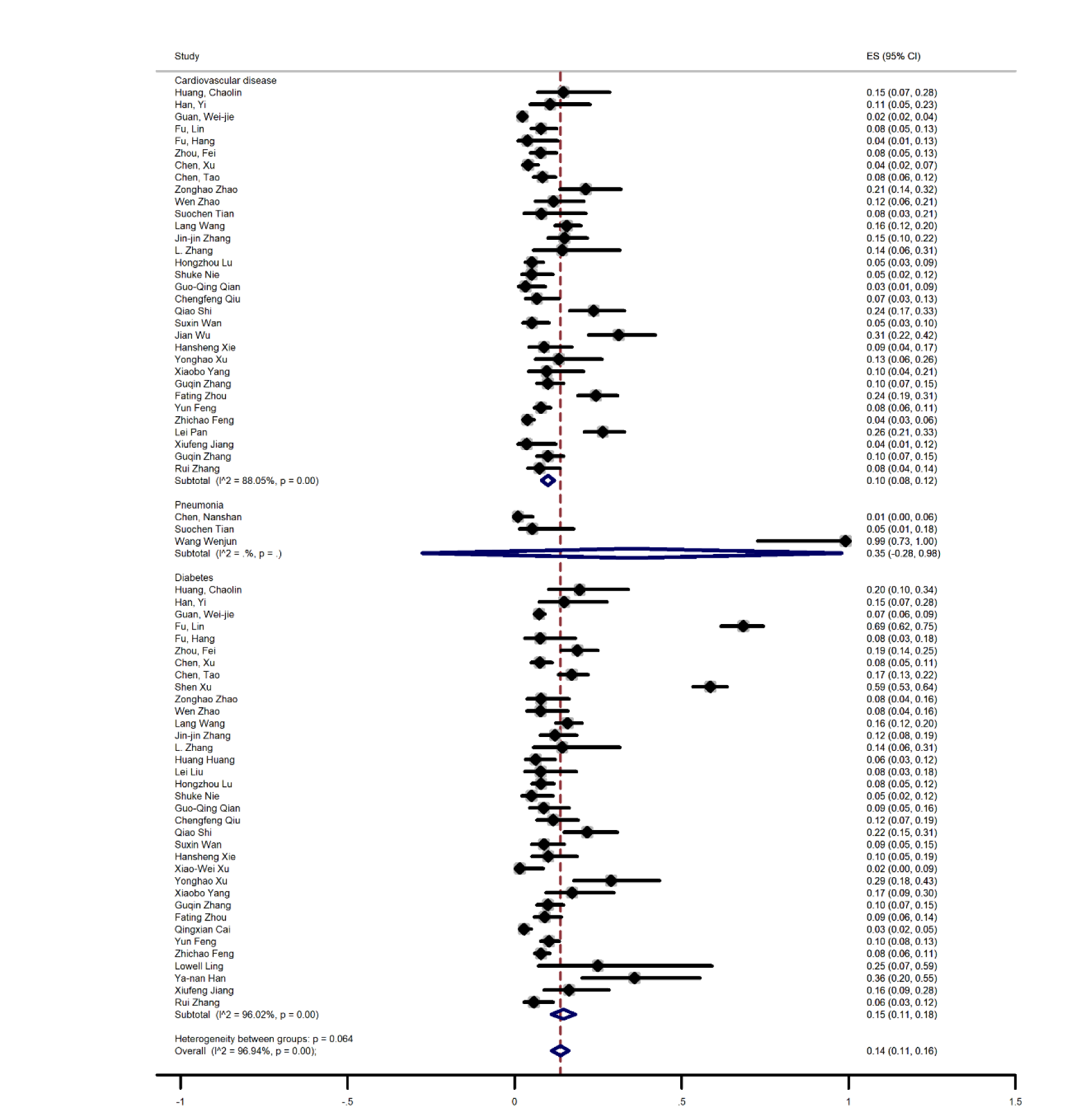
Meta-analysis of prevalence of comorbidities of all patients (A)

**Figure 8.**
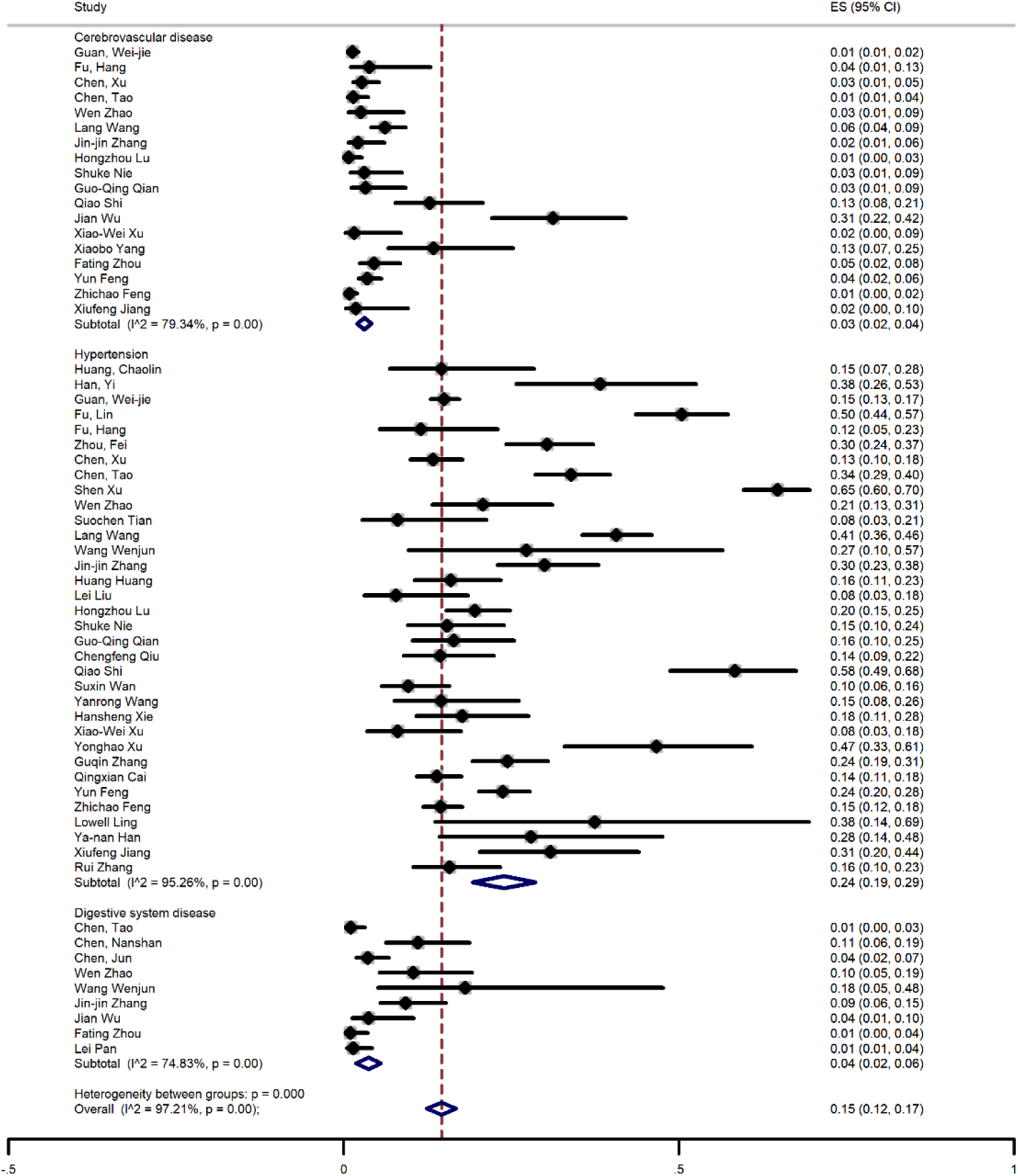
Meta-analysis of prevalence of comorbidities of all patients (B)

**Table 2.** Summarized Pooled values of Considered findings

| Variable | All patients |  |  | Severe |  |  | Non-severe |  |  |
| --- | --- | --- | --- | --- | --- | --- | --- | --- | --- |
|  | N of study | I-squared | ES% (95%CI) | N of study | I-squared | ES% (95%CI) | N of study | I-squared | ES% (95%CI) |
| **Clinical Outcomes** | - | - | - | - | - | - | - | - | - |
| Discharged | 32 | 99.86 | 50(35-65) | 12 | 98.07 | 29(21-37) | 7 | 99.94 | 43(1-84) |
| Death | 29 | 99.69 | 16(1-31) | 14 | 99.87 | 50(39-60) | 4 | 43.21 | 0(0-1) |
| Hospitalization | 23 | 99.77 | 80 (72-88) | - | - | - | - | - | - |
| ICU admission | 32 | 99.93 | 27(9-44) | - | - | - | - | - | - |
| **Laboratory findings** | - | - | - | - | - | - | - | - | - |
| Increase in ALT | 20 | 91.30 | 17(13-21) | 10 | 97.47 | 38 (18-58) | 6 | 97.09 | 30 (16-44) |
| Increase in AST | 19 | 92.59 | 18(14-23) | 11 | 94.03 | 48(34-63) | 8 | 90.79 | 21(13-29) |
| Increase in total bilirubin | 8 | 90.69 | 12(7-17) | 4 | 66.67 | 17 (9-25) | 2 | - | 10 (7-12) |
| Decrease in albumin | 6 | 99.87 | 44(0-88) | 3 | - | 36 (4-67) | - | - | - |
| Increase in C-Reactive Protein | 18 | 96.66 | 53(43-62) | 10 | 96.07 | 78 (69-88) | 7 | 98.57 | 55 (36-73) |
| Increase in LDH | 18 | 98.23 | 36(25-47) | 9 | 97.36 | 75(62-87) | 4 | 71.85 | 39(28-49) |
| Increase in ESR | 9 | 91.90 | 57(44-70) | - | - | - | - | - | - |
| Increase in D-dimer | 17 | 97.97 | 35(24-47) | 9 | 94.70 | 79 (70-89) | 6 | 96.07 | 28 (14-41) |
| **Comorbidities** | - | - | - | - | - | - | - | - | - |
| Hypertension | 34 | 95.26 | 24(19-29) | 23 | 85.64 | 38(31-45) | 15 | 88.58 | 13 (9-17) |
| Diabetes | 35 | 96.02 | 15(11-18) | 22 | 97.85 | 26(11-40) | 12 | 89.48 | 7 (4-10) |
| Cardiovascular disease | 32 | 88.05 | 10 (8-12) | 20 | 76.78 | 16(12-20) | 10 | 52.24 | 4(3-6) |
| Respiratory system disease | 26 | 62.40 | 3(2-4) | 15 | 60.21 | 6 (4-9) | - | - | - |
| Chronic liver disease | 25 | 75.59 | 3 (2-4) | 16 | 74.89 | 6 (3-9) | 10 | 81.36 | 4(1-6) |
| Pneumonia | 3 | - | 35 (-28-98) | - | - | - | - | - | - |
| Cerebrovascular disease | 18 | 79.34 | 3(2-4) | 12 | 64.03 | 7(4-9) | 6 | 0.97 | 1(1-2) |
| Digestive system disease | 9 | 74.83 | 4 (2-6) | 2 | - | 12 (5-19) | 2 | - | 8 (3-12) |
| Endocrine system disease | 7 | 78.60 | 8 (4-12) | - | - | - | - | - | - |
| Chronic kidney disease | 19 | 51.48 | 1(1-2) | 14 | 0 | 3 (2-4) | 5 | 0 | 0 (0-1) |
| CNS disorders | 4 | 0 | 1 (0-3) | - | - | - | - | - | - |
| Immunodeficiency | 7 | 51.54 | 1(0-1) | 3 | - | 7 (-2-17) | 5 | 0 | 0 (0-1) |
| Malignancy | 28 | 99.64 | 6 (1-11) | 12 | 21.94 | 3 ( 2-5) | 8 | 17.18 | 1 (1-2) |
| **Complications** |  |  |  |  |  |  |  |  |  |
| Liver toxicity | 13 | 95.35 | 20 (14-26) | 7 | 97.23 | 41 (19-62) | 2 | - | 17 (13-20) |
| Acute kidney injury | 6 | 84.51 | 8 (4-13) | 5 | 68.57 | 7 (2-13) | 3 | - | 2 (0-4) |
| Shock | 6 | 70.94 | 3 (1-5) | 3 | - | 11 (-3-24) | - | - | - |
| Acute respiratory distress syndrome | 6 | 95.69 | 28 (15-40) | 5 | 97.68 | 67 (44-90) | - | - | - |

Other disorders prevalence was as follows: Cerebrovascular disease 3% (95% Cl, 2-4), digestive system disease 4% (95% Cl, 2-6), chronic liver disease 3% (95% Cl, 2-4), malignancy 7% (95% Cl, 1-12), respiratory system disease 3% (95% Cl, 2-4), chronic kidney disease 1% (95% Cl, 1-2), endocrine system disease 8% (95% Cl, 4-12), immunodeficiency 1% (95% Cl, 0-1) and CNS disease 1% (95% Cl, 0-3) respectively (Fig.7, Fig.8, Fig.9, Fig.10 and Table 2). Pneumonia, acute respiratory distress syndrome (ARDS), acute kidney injury, and shock after infecting with COVID-19 have been investigated in 3, 6, 6, and 6 studies, respectively. Meta-analysis showed the prevalence of 35% (95% Cl, −28-98), 28% (95% Cl, 15-40), 8% (95% Cl, 4-13) and 3% (95% Cl, 1-5) respectively (Fig5, Fig.10 and Table 2). Also, liver toxicity as a complication of COVID-19 was reported in 13 studies with the prevalence of (20%, 95% Cl, 14-26).(Fig.5)

**Figure 9.**
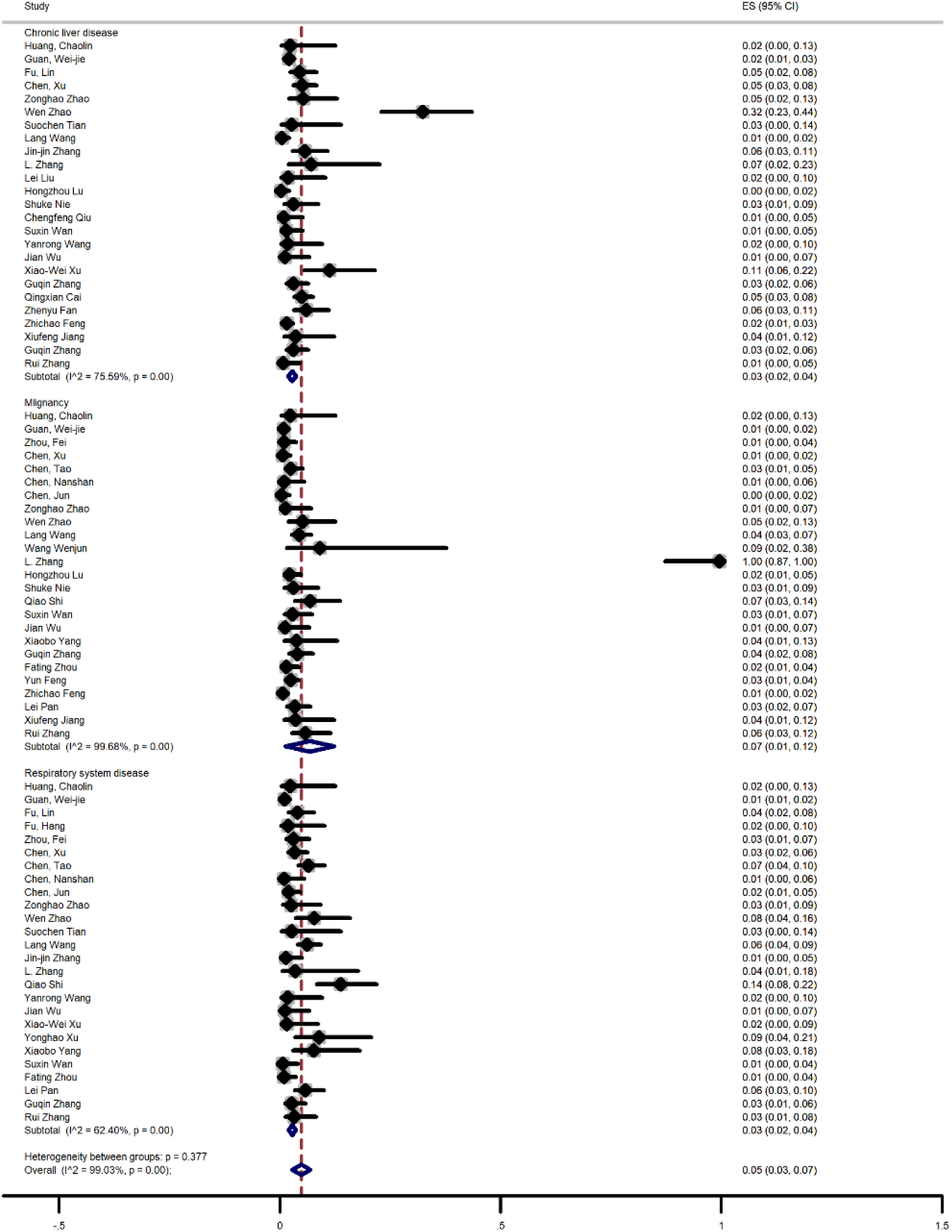
Meta-analysis of prevalence of comorbidities of all patients (C)

**Figure 10.**
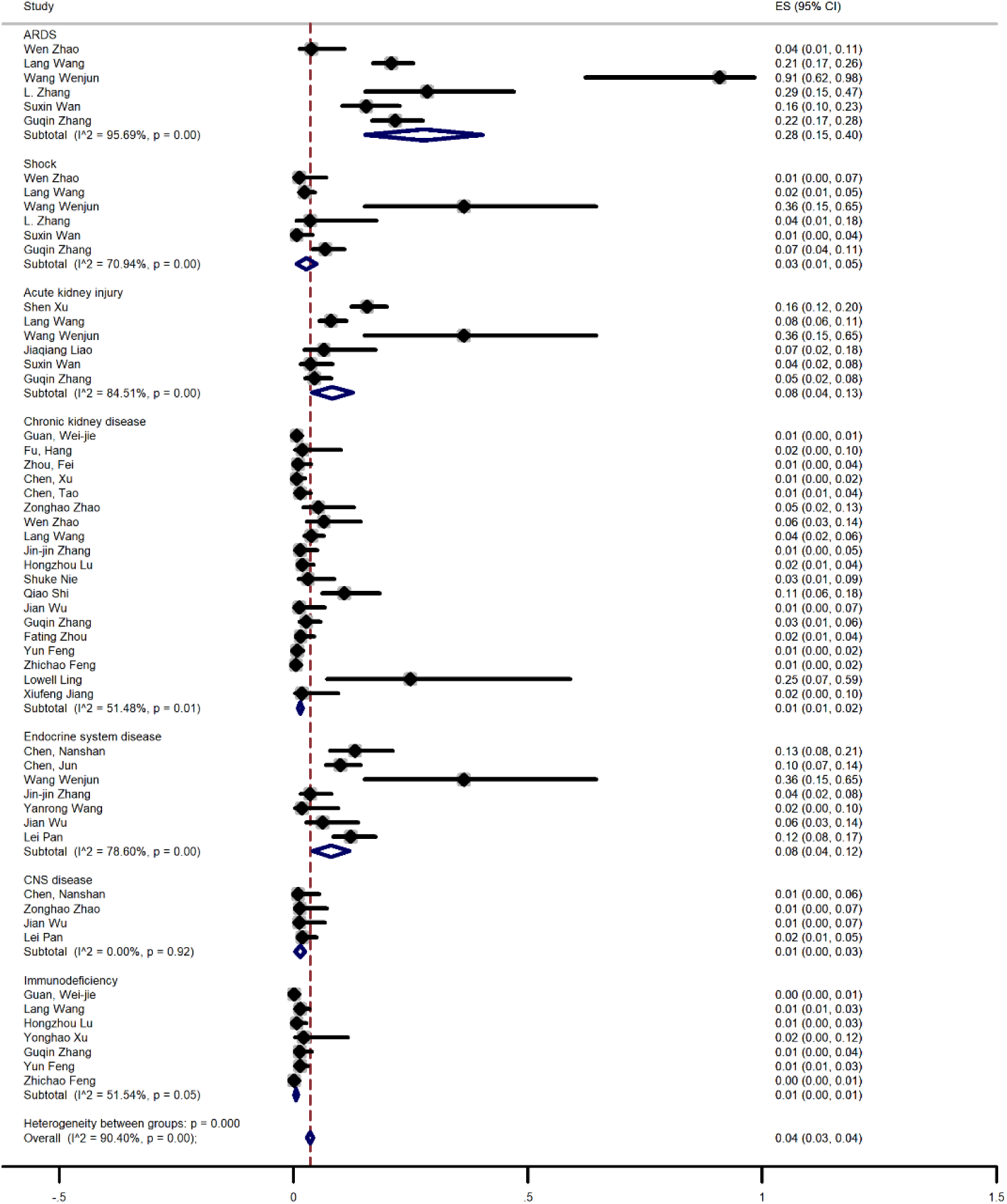
Meta-analysis of prevalence of comorbidities of all patients (D)

### Clinical outcomes of patients based on the severity

The meta-analysis showed that the prevalence of mortality among non-severe patients was 0% (95% Cl, 0-1), whereas 50% (95% Cl, 39-60) in severe patients. Furthermore, the prevalence of discharged patients among non-severe cases were 43% (95% Cl, 1-84), which were 29% (95% Cl, 21-37) in severe cases (Supplementary File and Table 2).

### Laboratory findings based on the severity of the disease

In this systematic review, we analyzed the subgroups based on the severity of the disease for finding the source of heterogeneity. The results are as follows:

The prevalence of increased liver enzymes among the patients, including ALT and AST were 30% and 21% in non-severe patients, respectively, which were 38% and 48% in severe patients (Supplementary File and Table 2).

The meta-analysis showed that the prevalence of increasing other laboratory findings like CRP, LDH, D-dimer, and Bilirubin were 55%, 39%, 28%, and 10% in non-severe patients respectively, which were 78%, 75%, 79% and 17% in sever patients. Also, the prevalence of decreasing albumin in severe patients was 36%, and the prevalence of liver toxicity among non-sever patients was 17%, whereas 41% in severe patients. (Supplementary File and Table 2).

### Comorbidities based on the severity of the disease

The prevalence of comorbidities such as hypertension, diabetes, and cardiovascular disease was 13%, 7%, and 4% in non-severe patients, respectively. However, these statistics were 38%, 26%, and 16%, respectively (Supplementary File and Table 2).

The prevalence of other underlying diseases like malignancy, digestive system disease, chronic liver disease, immunodeficiency, chronic kidney disease, and cerebrovascular disease were 1%, 8%, 4%, 0%, 0% and 1% in non-severe patients respectively whereas in severe patients were 3%, 12%, 6%, 7%, 3% and 7% (Supplementary File and Table 2).

## Discussion

The novel coronavirus 2019, which named SARS-Cov-2, shares about 80% of the genetic sequence with SARS-CoV, which they also have the same cell entry receptor (23). Based on studies was performed by Zhao et al., it is demonstrated that 83% of ACE2-expressing cells were alveolar epithelial type II cells (AECII), suggesting that these cells can serve as a reservoir for viral invasion (24). Extrapulmonary sites such as the heart (25), kidney (26), endothelium (27), intestine (28), and liver (29) also expressed the ACE2 receptor. During recent studies, it is revealed that SARS-CoV-2 can bind to the cholangiocyte ACE2 receptors leading to cholangiocyte dysfunction and, through the induction of a systematic inflammatory response, causes liver injury (30). Also, Through liver biopsy assessing of the deceased case of COVID-19, moderate microvesicular steatosis, a common finding in sepsis (31), and mild lobular and portal activity were observed, which confirming that this injury could occur; as a result SARS-CoV-2 infection (32). In another theory, it is postulated that viral-induced cytotoxic T cells and imbalanced innate immune system lead to collateral liver damage, indicating the association between abnormal liver markers and COVID-19 disease severity (33, 34).

Recent studies on SARS clinical features have revealed that the liver enzyme abnormalities are common but not as the prominent feature of this illness (35). Through the recent pandemic of SARS-CoV-2, liver dysfunction in many varieties has been reported in many cases. In this systematic review and meta-analyses, we aimed to evaluate the prognoses of liver damage. Liver test abnormality at the admission time can be used as a predictor for the severity of the disease (36). In this case, it is assumed that enhanced end-expiratory pressure by increasing right atrial pressure lead to hepatic congestion and blocked venous return. So many patients admitted to the hospital are with liver abnormalities in the absence of mechanical ventilation (33).

Acute liver injury caused by SARS-CoV-2 is more common in patients with poor prognosis who had significantly higher risks of developing severe pneumonia, and Male elderly COVID-19 patients with diabetes and lymphopenia. In the early stages, it could enhance the mortality risk in COVID-19 patients. Also, it is reported that hepatic functional indexes of two-third COVID-19 patients remain poor in two weeks after discharge as the consequence of Compassionate drug usage during hospitalization due to lacking specific drug for SARS-CoV-2 (37, 38).

In the current study, trough analyses, results showed significantly higher liver toxicity in sever patients than non-sever patients. The liver toxicity (hepatotoxicity) is a common adverse event that could occur during clinical practice because of the leading systemic toxicity of drugs and chemicals (39). Many drugs used for COVID-19 patients also could damage the liver and among them lopinavir/ritonavir associated with seven times higher odds of liver injury, which have been reported to cause liver damage and affect liver tests (36, 40). Other drugs that have been used for the treatment of COVID-19 patients, such as antibiotics, antivirals, and steroids, could potentially damage the liver, but still not being evident (41). However, for hospitalized patients, more attention should be paid to drug-induced liver damage. All to gather Liver enzymes elevation could be due to drugs used for treatment, sepsis, and shock (36).

In this systematic and meta-analyses review, we observed a higher amount of ALT, AST, LDH, D-Dimer, CRP, and total Bilirubin (TBIL) with slightly lower albumin in sever group compared to non-sever groups.

Based on previous studies on the ACE2 receptor, it is reported that slight amounts of these receptors were expressed in human hepatocyte (27, 42), indicating the non-significant influence of liver function by SARS-CoV-2 infection in non-sever and mild cases (43, 44).

Recent studies have reported the incidence of liver damage in severe cases of COVID-19 with mostly higher ALT, AST, LDH, CRP, D-dimer, and total bilirubin amount and lower albumin (36, 37, 43-45). Also, it has been mentioned that D-dimer, C-reactive protein, and AST higher levels were associated with COVID-19 severity (44). Moreover, through a study on deceased cases of COVID-19 with liver abnormalities, it is reported that in patients with severe complications, ALT, AST, and TBIL were higher than the standard limit. Furthermore, total albumin was also reported lower than the limited upper unit (46). In addition, in other studies, lower albumin levels in severe cases also have been reported (44, 47). Hypoalbuminemia could occur as a result of inadequate nutrition intake and overconsumption during hospitalization (43).

Based on the result of this study, patients who have the severe condition are 5.54, 4.22, 4.96, 4.13 and 4.34 times more likely to have elevated CRP, ALT, AST, LDH, D-dimer enzymes retrospectively. However, the chance of developing liver toxicity as a complication of COVID-19 was 1.76 times more in severe patients. Also, in a similar study, elevated serum levels of AST, ALT, and total Bilirubin, and lower levels of Albumin were observed in severe cases.

Their meta-analyses for AST (95% CI = 5.97 to 11.71, I^2^ = 73.4%), ALT (95% CI = 4.77 to 9.93, I^2^= 57.2%), TBIL (95% CI = 1.24 to 3.36, I^2^ = 68.8%), and Albumin (95% CI = −6.20 to −2.28, I^2^= 95.7%) (21) results were difference with us where AST(95% Cl, 14-23, I^2^= 94.03%), ALT (95% Cl, 13-21, I^2^= 97.47%), and TBIL (95% Cl, 7-17, I^2^= 66.67%), (95% Cl, 0-88, I^2^=0%) in severe cases accompanied with better results except of Albumin due to higher included studies.

In another systematic review and meta-analyses survey on liver function during COVID-19, it is reported that elevated AST(95% CI 13.6 to 16.5) and ALT(95% CI 13.6 to 16.4) were seen in 15% of patients; also, increases in TBIL was seen in 16.7% (95% CI 15.0 to 18.5) of patients. Theses upper than normal limit increases were attributed to drug or viral induced hepatic injury. Drug-induced hepatotoxicity also was reported with Remdesivir and Favipiravir. Notably, Lopinavir/ritonavir liver injury had not been evident in COVID-19 patients, and liver injury with Chloroquine and Hydroxychloroquine reported rare. They not specifically compared these increases in non-severe than severe patients (48).

Some studies also reported that LDH was higher in COVID-19 patients with severe pneumonia. LDH is an essential element in glucose metabolism and expressed widely through organ body tissues, especially in myocardial and liver cells. LDH would release within cells upon the cytoplasmic membrane damage (34). Based on previous studies on SARS and MERS, the elevated levels of LDH also were reported. So it can be concluded that LDH independently could act as a risk factor with poor clinical outcomes and raised the alarm for further analyses (45, 49). The higher levels of LDH maybe is due to the broad expression of ACE2 receptors in cardiac blood vessels (27, 50). In another theory, the increased amount of LDH was attributed to myositis, which resulted from virus infection (33). Moreover, its amounts would be increased as a cause of hepatocyte injuries because of the coexisting of ACE2 receptors in hepatocytes. So it’s lightening this fact that liver and cardiac damage could occur as a direct effect of SARS-Cov-2 on targeted organ damage (34). Thus elevated LDH enzyme in sever cases might occur as a consequence of directed hepatic or extrahepatic damage.

Like SARS-CoV, SARS-CoV-2 could affect lymphocytes, especially T lymphocytes (34). Patients with damaged T cells are more vulnerable to infections and their risk increase in patients with severe illness. The CRP in sever patients had higher levels than non-sever, and the CRP level above 100mg/dl could be an alarm for bacterial infection. So higher CRP might indicate the risk of other infections (mostly opportunistic infections), which could affect the liver or lead to hepatitis, and act as a prognostic factor (51-54). On the other hand, cytokine factors that were marked by higher concentrations of CRP, ESR, ferritin, and hs-CRP are associated with the severity of COVID-19. As a result, higher amounts of cytokine factors in blood serum might suggest a pro-inflammatory factor storm (55).

Another severe complication that occurred in COVID-19 patients is sepsis, and it could be with some clinical and labratorical manifestations. Through labratorical analyses, hyperbilirubinemia, acidosis, high lactate, coagulopathy, and thrombocytopenia were mostly seen in COVID-19 patients in ICU (56, 57). Also, as mentioned above, sepsis is one of the liver injury causes during infection with SARS-Cov-2.

Trough abnormal laboratory findings in severely affected patients, aberrant coagulation has been reported (58). Also, based on recent studies, it is revealed that COVID-19 is associated with the classical syndrome named disseminated intravascular coagulation (59) and the subsequent consumption coagulopathy (60). The D-dimer is a fibrin degradation product which highly increases during DIC (61). Besides, The elevation in D-dimer levels could occur as a result of cirrhosis and also are increased gradually as the degree of liver dysfunction increases in severity (62, 63). All to gather higher levels of D-dimer with hepatic or extrahepatic origin could be act as a prognostic factor for COVID-19.

Nevertheless, In a study, it is reported that liver indexes such as ALT, AST, TBIL, ALP, ALB, GLB, INR, LDH, and CRP hadn’t independently any association with the severity of COVID-19 (43).

All to gather in this systematic review and meta-analyses, abnormal liver markers where different in sever group than the non-sever group. Higher ALT, AST, LDH, CRP, D-dimer, and total bilirubin amount and lower albumin altogether but not independently could be a prognostic factor for COVID-19 patients.

The present study has some limitations. All of the studies have been conducted in China, whereas covid19 is pandemic now. Interpretation of our meta-analysis findings might be limited by the small sample size. limitations on the information provided in studies constrained subgroup analysis.

## Conclusion

The liver because of ACE2 expression in this site is prone to injury as a result of infection with SARS-CoV-2. Through our meta-analyzes elevation of some liver, markers were higher in sever group when compared to the non-sever group. Also, it is notable that Albumin was significantly reduced in severe compared to non-sever patients. Besides some medications were used to treatment of COVID-19 patients could be toxic for the liver. Through results, we observed significantly considerable numbers of drug-induced liver toxicity in sever group than the non-sever group. All to gather in the current study, we assumed that abnormal liver markers could act as a prognostic factor for a better survey of COVID-19.

## Data Availability

The data that support the findings of this study are openly available in data bases mentioned in the search strategy.

## Acknowledgment

The authors would like to thank the Student Research Committee of Mazandaran University of Medical Sciences for supporting this project. This study was approved in the Ethical Review Committee of the Mazandaran University of Medical Science with ethical code IR.MAZUMS.REC.1399.049.

## Supporting Information Legend

Supplementary Figure 1. Meta-analysis of prevalence of clinical outcome of non-severe patients.

Supplementary Figure 2. Meta-analysis of prevalence of clinical outcome of severe patients.

Supplementary Figure 3. Meta-analysis of prevalence of laboratory findings of non-severe patients.

Supplementary Figure 4. Meta-analysis of prevalence of laboratory findings of severe patients.

Supplementary Figure 5. Meta-analysis of prevalence of comorbidities of non-severe patients (A)

Supplementary Figure 6. Meta-analysis of prevalence of comorbidities of non-severe patients (B)

Supplementary Figure 7. Meta-analysis of prevalence of comorbidities of severe patients (A)

Supplementary Figure 8. Meta-analysis of prevalence of comorbidities of severe patients (B)

Supplementary Figure 9. Meta-analysis of prevalence of comorbidities of severe patients (C)

